# Novel subgroups of type 2 diabetes based on multi-Omics profiling: an IMI-RHAPSODY Study

**DOI:** 10.1101/2022.09.03.22279563

**Authors:** Shiying Li, Iulian Dragan, Chun Ho Fung, Dmitry Kuznetsov, Michael K. Hansen, Joline W.J. Beulens, Leen M. ’t Hart, Roderick C. Slieker, Louise A. Donnelly, Mathias J. Gerl, Christian Klose, Florence Mehl, Kai Simons, Petra JM Elders, Ewan R. Pearson, Guy A. Rutter, Mark Ibberson

## Abstract

Type 2 diabetes is a complex, multifactorial disease with varying presentation and underlying pathophysiology. Recent studies using data-driven cluster analysis have led to a stratification of type 2 diabetes into novel subgroups based on six clinical measurements. Whether these subgroups truly correspond to the underlying phenotypic differences is nevertheless unclear. Here, we apply an unsupervised, data-driven clustering method (Similarity Network Fusion) to characterize type 2 diabetes in two independent cohorts involving 1,134 subjects in total based on integrated plasma lipidomics and peptidomics data without pre-selection. Logistic regression was then used to explore clustering based on ≥ 180 circulating lipids and 1,195 protein biomarkers, alongside clinical signatures. Two subgroups were identified, one of which associated with elevated C-peptide levels, diabetic complications and more severe insulin resistance compared to the other. GWAS analysis against 403 type 2 diabetes risk variants revealed associations of several SNPs with clusters and altered molecular profiles. We thus demonstrate that heterogeneity in type 2 diabetes can be captured by circulating omics alone using an unsupervised bottom-up approach. Such multiomics signatures could reflect pathological mechanisms underlying type 2 diabetes and thus may help inform on precision medicine approaches to disease management.

## Introduction

Type 2 diabetes is global problem of increasing proportions and a substantial threat to human health (1). Recently, Ahlqvist et al. (2) proposed an approach to sub-classifying patients with type 2 diabetes into four different subgroups by *K-means* clustering using six clinical measurements: age at diagnosis, Body Mass Index (BMI), Glutamic acid decarboxylase (GAD) autoantibodies (GADA), HOMA2-B (beta-cell function), HOMA2-IR (insulin resistance) and HbA1c. Since then, we (3) and others (4,5) have applied a similar clustering method with independent cohorts and obtained similar results. These studies suggest the new classification system facilitates meaningful sample subgroup discovery across different populations. However, the value of this stratification approach has been questioned by some researchers (6,7) in terms of its stability and utility. Importantly, it is still debatable whether type 2 diabetes patients with different underlying aetiologies are represented faithfully, by placing them into subsets, based solely on a limited number of clinical features. Indeed, others (8) have proposed that positioning individuals within a multi-dimensional, and potentially fluid, continuum of variables may provide a more useful prognostic strategy.

An alternative clustering approach, Similarity Network Fusion (SNF) (9) combines multiple levels of data from the same patients into a similarity network, enabling exploratory multidimensional data-driven clustering. First, patient similarity networks are generated for each data level independently then merged into a single global network through an iterative process. Clustering can then be performed on this network using a graph-based algorithm. As opposed to clustering patients based on a limited number of pre-selected clinical features, this unsupervised bottom-up approach can leverage much larger biological data sets (“omics”), and holds the promise of uncovering new insights, which may be missed by models based on single data types, limited clinical features or supervised approaches.

In the present study, we performed distributed analysis of two independent patient cohorts to explore the stratification of type 2 diabetes based on combined plasma lipidomics and peptidomics using a federated database system. In addition, we explore the genetic factors predisposing to membership in these groups and the potential correlations with patient molecular profiles.

## Methods

### Study populations

We used data from two type 2 diabetes cohorts: Hoorn Diabetes Care System (DCS) and Genetics of Diabetes Audit and Research in Tayside Scotland (GoDARTS). The DCS cohort recruits almost all type 2 diabetes patients from 103 GPs in the West-Friesland region of the Netherlands. This prospective, regional cohort study started in 1998 and by the time of 2017, it holds 12,673 type 2 diabetes patients with a median of 0.7 years (IQR 0.2-3.7) after diagnosis (10).

Between 1996 and 2015, GoDARTS recruited 10149 type 2 diabtes patients from the Tayside region of Scotland. Patients in the GoDARTS cohort were not necessarily recruited at the time of diagnosis (11).

Lipidomics and peptidomics are available for a subset of type 2 diabetes patients in both DCS and GoDARTS cohorts. These data were generated as part of the IMI2 RHAPSODY project (3). The sample availability for omics was within 2 years of diagnosis. Of note, individuals were selected without taking into consideration pre-cluster assignments.

### Measurements

Data from both cohorts were used with informed consent obtained through the relevant local ethical committees. In DCS, all blood measurements are performed in fasted individuals. In contrast, all measurements in GoDARTS are performed in the non-fasted state. In DCS, Haemoglobin A1c was measured based on the turbidimetric inhibition immunoassay for haemolysed whole EDTA blood (Cobas c501, Roche Diagnostics, Mannheim, Germany). The levels of triglycerides, total cholesterol and HDL cholesterol were measured enzymatically (Cobas c501, Roche Diagnostics) (10). In DCS and GoDARTS, C-peptide was measured on a DiaSorin Liaison (DiaSorin, Saluggia, Italy). Plasma lipids were determined using a QExactive mass spectrometer (Thermo Scientific). Plasma protein levels were measured on the SomaLogic SOMAscan platform (Boulder, Colorado, USA). For more details refer to (3).

### Cluster analysis

A federated database of type 2 diabetes cohorts including DCS and GoDARTS has previously been set up as part of the RHAPSODY project. This system enables statistical and machine learning analysis to be performed on cohort data at a distance without any disclosure of sensitive data. The federated database system was interrogated using the R statistical programming language (version 4.0.4). In both DCS and GoDARTS cohorts, patients with complete lipidomics and peptidomics (589 and 545 patients, DCS and GoDARTS, respectively) were used for clustering and subsequent statistical analysis. Lipidomics and peptidomics values were centered to a mean value of 0 and an SD of 1 in each cohort using the *scale* function in R. Euclidean distances between each pair of patients were then calculated based on the normalized lipidomics or peptidomics data by using *dist2* function from *dssSNF* (*dsSwissKnifeClient* package) (12). Patient similarity matrices were generated from the Euclidean distance matrices with parameters of K (the number of nearest neighbours) equal to 20 and hyperparameter alpha equal to 5. The patient similarity matrices for lipidomics and peptidomics were then fused using the *SNF* function within *SNFtool* package (13) with T (number of iterations) equal to 20. In both DCS and GoDARTS, men and women were clustered together without pre-separation. Two-step clustering was then performed, in which the optimal number of clusters is estimated in the first step using the silhouette method and patient assignments made in the second step. Silhouette widths of fused cluster patients were calculated from the similarity matrices using the *Silhouette_SimilarityMatrix* function (*CancerSubtypes* package) (14).

SNF models were validated by bootstrapping tests (n=1000 iterations) that compared model classification to models using randomized Euclidean distance matrices. Euclidean distance matrices were randomized using the function *randomizeMatrix* within the *picante* package (15). For each simulated model, the same parameters were used and the number of clusters is set to correspond to the number of clusters in the study model. The significance of each cluster (P-value <=0.05) was calculated by assessing the frequency of achieving a model with an equal or greater mean local cluster coefficient for randomized data. Local cluster coefficient calculation was done with unweighted, undirected adjacency matrices using the *transitivity* function in the *igraph* package (16). The adjacency matrices were generated from the similarity matrices with the top 5% similarity values set as 1 and the rest as 0.

The possibility of using clinical measurements to replicate our multiomics clustering results was assessed in a logistic regression model with pseudo R square as the recorded results. One of the common interpretations of pseudo R square is the indication of the improvement to which the model parameters improve upon the prediction of the null model. In a logistic regression model, we treated the patient’s assignment results as the dependent variable (cluster 1:1; cluster 2:0) with six clinical measurements: age, BMI, C-peptide, LDL cholesterol, HDL cholesterol, Triglycerides act as univariate or together as covariates. Pseudo R square=1-(deviation/ null deviation).

### Statistical analysis

Logistic regression modelling was performed by using the *ds*.*glm* function within the *dsBaseClient* package to assess the association level of each molecule (lipidomics and peptidomics), clinical measurements and type 2 diabetes SNPs (17) with the multiomics clusters. The cluster classification was treated as a dependent categorical variable with age, gender and BMI acting as covariates. Subsequently, for lipids, peptides and clinical measurements, each cluster was subset for the strongly associated (P-value <=0.05) features and the mean values of each group of features in each cluster were calculated. Mean values were used since, to protect the patient’s identity, individual-level data cannot be downloaded from the federated database system. However, it is possible to display a mean value if it is derived from 5 or more patients. Several mean values for each feature from 5 or more patients were calculated based on the default patients’ order in the remote server for each cluster. The feature values were normalized. Hierarchical clustering was then performed, and the results were visualized as heatmaps using the R package *gplots* (18).

For each patient in the database, the clinical cluster assignments were defined previously, similar to (2), but with five clinical variables: age, BMI, HbA1c, high-density lipoprotein (HDL) and C-peptide (3). Our clusters were compared with these clinical clusters by calculating the hypergeometric overlap p-value *(phyper* function in R).

Cox regression was performed using the *dssCoxph* function in the *dssSwissKnifeClient* package. Time to insulin requirement was defined as the length of time from diagnosis until an individual started insulin treatment for a period of more than six months, or alternatively as more than two independent HbA1c measurements greater than 69 mmol/mol (8.5%) at least three months apart and when ≥2 non-insulin glucose-lowering drugs were taken. Hazard ratios for time to insulin treatment requirement within the clusters were calculated using Cox regression with age, gender and BMI as covariates.

The associations between type 2 diabetes SNPs (17) and multiomics clusters or molecules with altered expression profiles were assessed using a linear regression model with age, BMI and gender as co-independent variables. The routinely used genome-wide common variant association threshold *p*⍰≤⍰5⍰×⍰10−8 was applied in this study (19) to assess significance.

All analyses were performed using the R statistical programming language (version 4.0.4). The Benjamini-Hochberg procedure was used to determine the false discovery rate correcting for multiple tests. Figures were produced using *gplots* (version 3.1.1), *ggplot2* (version 3.3.4) and *igraph* (version 1.2.6). Figure 1a was generated using *Biorender*.

**Figure 1.**
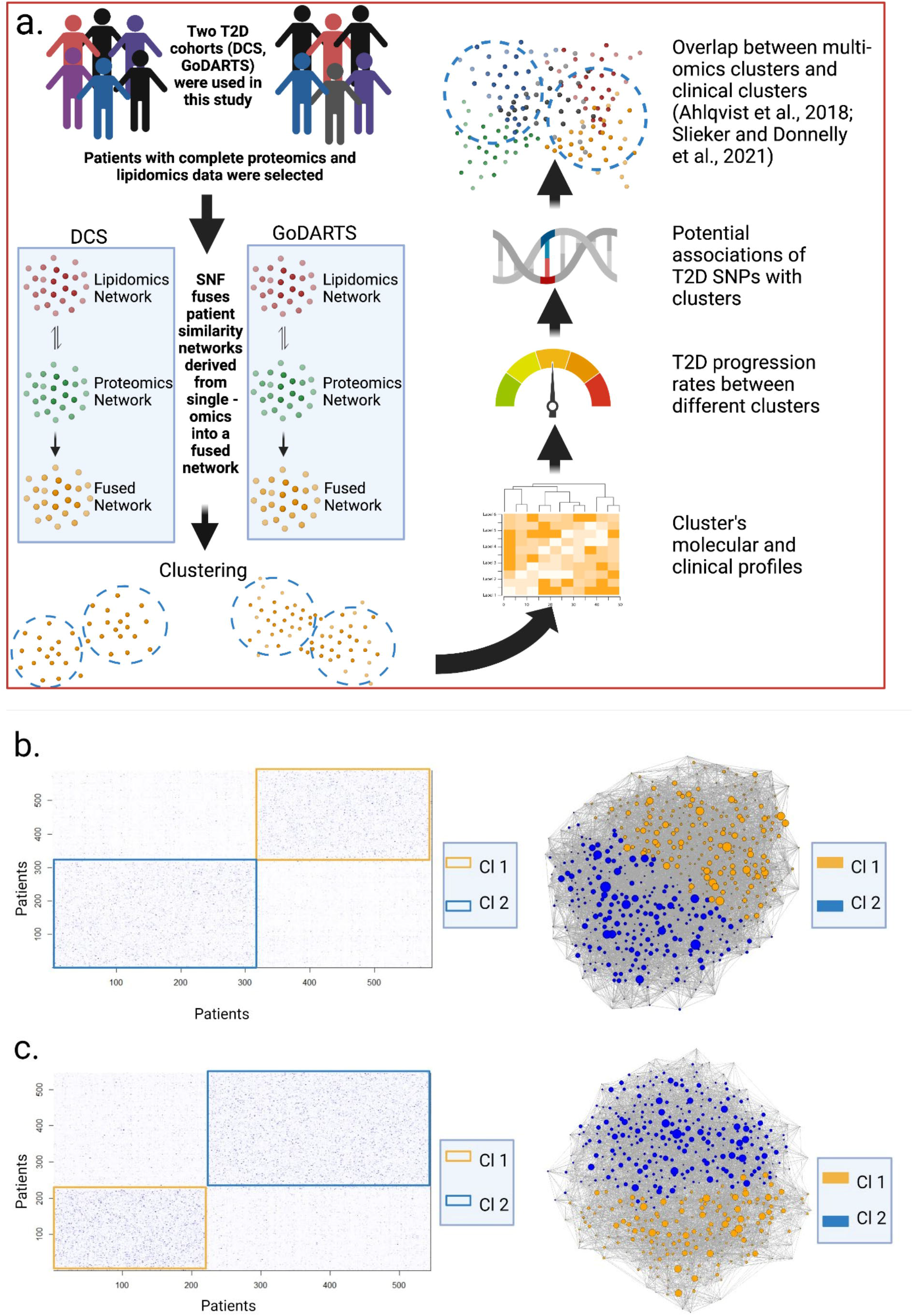
Experiment design and the molecular clusters displayed as both similarity matrices and patient network. (**b**,**c**) Molecular clusters were generated by integrated multiomics (lipidomics and peptidomics) data in both DCS (b) and GoDARTS (c), respectively. The similarity matrices were calculated based on Euclidean distances with parameters of K=20 (the number of shared neighbours of datapoint), alpha=0.5 (hyperparameter) and T=20 (number of iterations). Each point in the similarity matrices represents patient-to-patient similarity, the higher the colour intensity, the more similar the patients. Boxed regions represent the potential clusters. The patient networks were generated based on unweighted adjacency matrices. The nodes represent the patients, edge thickness reflects the strength of similarity, the size of the node represents the betweenness. Nodes were coloured based on their cluster assignments. DCS= Hoorn Diabetes Care System. GoDARTS=Genetics of Diabetes Audit and Research in Tayside Scotland.

### Data availability statement

Information about accessing the Rhapsody federated database can be found at: https://imi-rhapsody.eu/. Summary statistics of lipidomic and proteomic data will be available from an interactive Shiny dashboard, available upon publication.

## Results

### Unsupervised multi-Omics clustering defines two subgroups in independent cohorts

Similarity Network Fusion (SNF) was performed using plasma lipidomics and peptidomics data from a total of 1022 type 2 diabetes patients in two independent cohorts. 589 and 545 type 2 diabetes patients, respectively, from the DCS and GoDARTS cohorts with complete cases of plasma lipidomics and peptidomics measurements were selected. The characteristics of these two cohorts were generally comparable in terms of average age and BMI, with a majority of males (3). Individuals were clustered using SNF as described in Methods. Through silhouette analysis, we were able to demonstrate that clustering integrating both lipidomics and peptidomics outperforms either of the single -omics in terms of cluster assignment quality (Supplemental Figure 1). Based on a two-step clustering procedure, the optimal number of clusters was found to be two (Supplemental Figure 1). The significance of the clustering results was validated by bootstrapping (n=1000 iterations) against simulated datasets (Supplemental Figure 2). In the DCS cohort, cluster 1 comprised 46.5% of the patients and was characterized by a relatively high BMI value compared to cluster 2 (53.5%). In both GoDARTS (cluster 1:40.7%; cluster 2:59.3%) and DCS cohorts, patients have similar age, HbA1c values. Furthermore, in the DCS cohort, triglycerides, C-peptide and homeostatic model assessment (HOMA) 2, HOMA2-B and HOMA2-IR, which were calculated based on fasting blood glucose and fasting C-peptide concentration, showed a higher concentration in cluster 1 compared to cluster 2. In contrast, LDL cholesterol, HDL cholesterol and HOMA2-S show a higher concentration in cluster 2 (Supplemental Figure 3). Similarly, a higher concentration of LDL cholesterol and HDL cholesterol, and a lower concentration of triglycerides and C-peptide, were also observed in cluster 2 compared to cluster 1 in the GoDARTS cohort (Figure 2a; Supplemental Figure 3).

**Figure 2.**
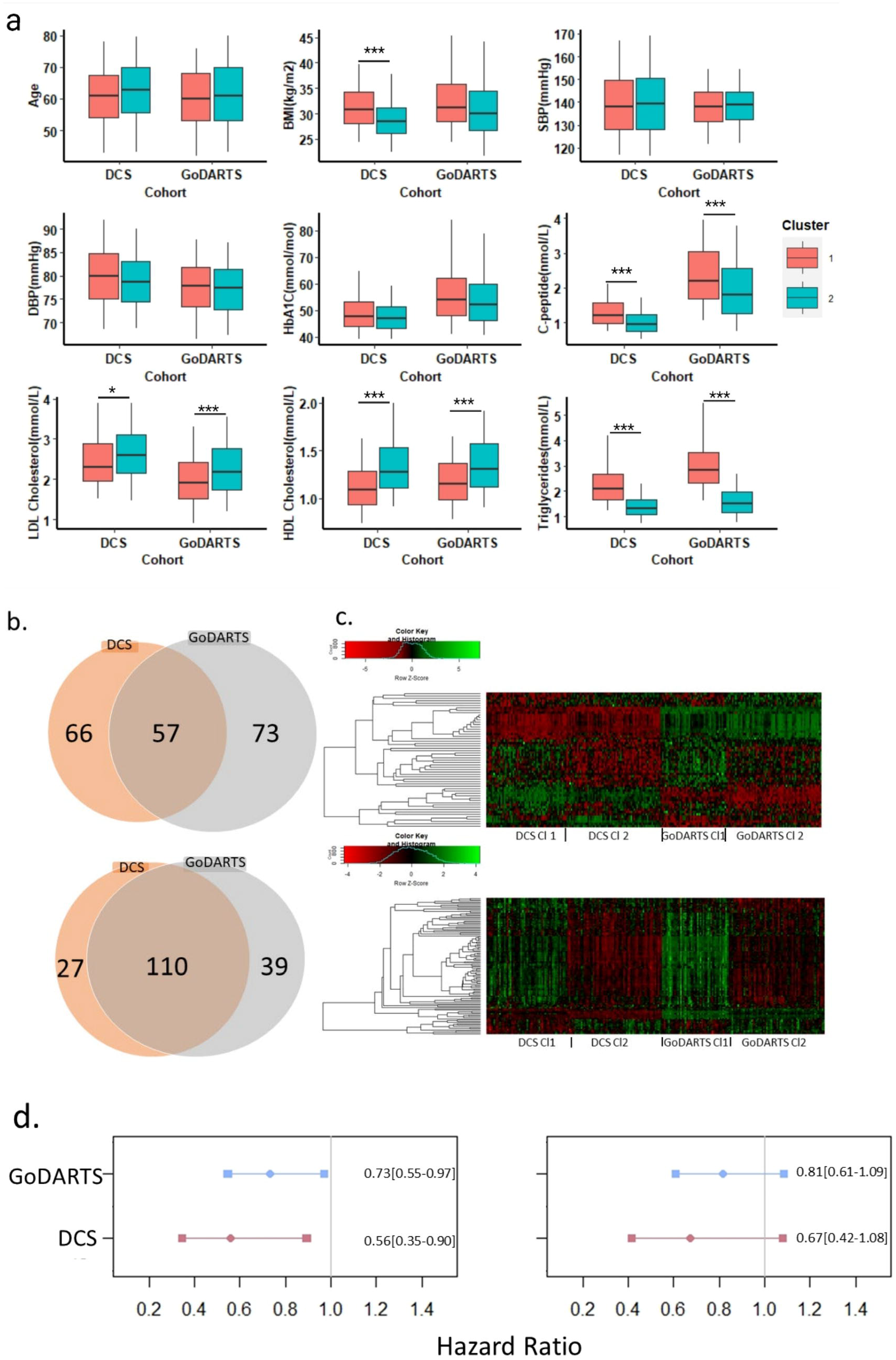
(a,b,c) Clinical characteristics and molecular profiles of multi-omics clusters in both DCS and GoDARTS clusters. (d) The hazard ratio for time to insulin requirement across clusters. Multi-omics clustering was done without separating the women and men. In each cohort, the statistical difference between cluster 1 and cluster 2 was determined by logistic regression with age, BMI and gender as covariates. Multi-analysed P-values were adjusted by the Benjamini-Hochberg procedure, and a false discovery rate (FDR)-adjusted P-value <=0.05 was considered significant. (**a**) Distributions of nine clinical measurements at baseline in both DCS and GoDARTS cohorts for each cluster. *:P-value<=0.05. **:P-value<=0.01. ***:P-value<=0.001. (**b**) Venn diagrams showing overlaps of significant (significantly different between clusters) proteins (upper) or lipids (lower) across DCS and GoDARTS cohorts. (**c**) The relative concentration heatmaps of common significant proteins (upper) or lipids (lower) between DCS and GoDARTS cohorts. Concentrations of each feature were first converted into the log(concentration) and then normalized in order to investigate the relative expression level of each feature between different clusters (Row Z-score). Heatmap cells were coloured in red/green to represent the relative expression level (red=low, green=high). The federated database does not allow to download data from individual samples in order to protect the patient’s identity. Each vertical line in the heatmap represents 5 different patients’ mean values for each feature. y-labels were hidden for a better presentation. Fully labelled versions are available in Supplemental Figure 5. (**d**) X-axis, hazard ratio, y-axis, database. Cox model with correction of gender (left) or with corrections for gender, age and BMI (right). The number in the panel represents the hazards with 95% confidence intervals. Multi-omics cluster 1:1 and Multi-omics cluster 2:2. Cox model gives the hazard ratio for the second group relative to the first group. The hazard ratio is not significant if the 95% confidence interval includes 1. DBP= Diastolic Blood Pressure. SBP= Systolic Blood Pressure. HbA1C= Haemoglobin A1C. DCS= Hoorn Diabetes Care System. GoDARTS=Genetics of Diabetes Audit and Research in Tayside Scotland.

**Figure 3.**
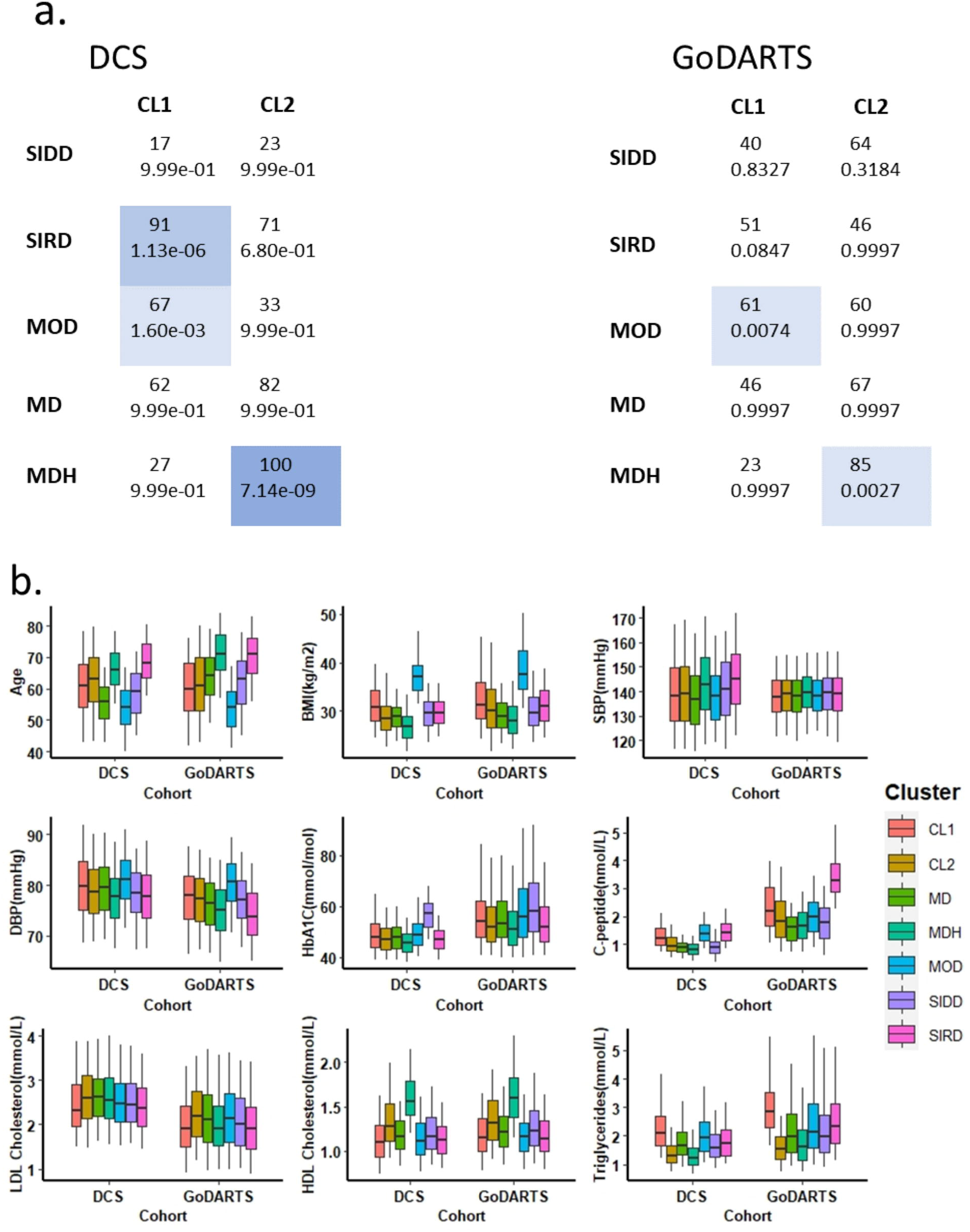
**(a)** Comparison of clustering in Slieker and Donnelly et al., 2021 versus clustering in the current study. The first number in each cell represents the number of individuals overlapping, the second number represents the hypergeometric overlap p-value. **(b)** Distributions of nine clinical measurements at baseline in both DCS and GoDARTS cohorts for multi-omics clusters and clinical clusters. CL1= multi-omics cluster #1. CL2= multi-omics cluster #2. MD= mild diabetes. MDH= mild diabetes with high HDL. MOD= mild obesity-related diabetes. SIDD= severely insulin deficient diabetes. SIRD= severely insulin resistant diabetes. SBP=systolic blood pressure. DBP=diastolic blood pressure. HbA1C=Hemoglobin A1C.

We compared type 2 diabetes progression rates between clusters in both cohorts using Cox Proportional hazard models. In both cohorts, type 2 diabetes progression rate showed a nominally significant difference in DCS and GoDARTS, respectively, between multi-omics cluster 1 and cluster 2 (*p*=0.0156 (unadjusted), DCS; *p*=0.0308 (unadjusted), GoDARTS; corrected for gender). Cluster 2 reduces the hazard ratio by 44% or 27% in DCS or GoDARTS, respectively (Figure 2d, left). Results were in the same direction but were less significant when additionally correcting for age and BMI (figure 2d, right).

We then investigated whether the multi-omics clusters could be replicated by using clinical measurements alone. Pseudo R square was used to assess the possibility of using six clinical features: age, BMI, LDL cholesterol, HDL cholesterol, C-peptide and triglycerides to predict the multi-omics clustering results in a logistic model for both cohorts (Supplemental Table 1). The highest pseudo R^2 scores are shown when all seven clinical measurements act as covariates with 29.9% and 46.9% improvement over random in DCS and GoDARTS, respectively. These results clearly indicate that the molecular clusters we observe are not merely reflecting differences in routine clinical measurements.

In summary, by combining plasma lipidomics and peptidomics data in a data-driven and unsupervised analysis, we were able to separate type 2 diabetes patients from two independent cohorts into two subgroups that appear to differ in diabetes-related clinical characteristics.

### Similar molecular signatures separate subgroups in both cohorts

We investigated the correlation of each plasma lipidomics or peptidomics feature with the clustering results using logistic regression models adjusting for gender, BMI and age. In DCS, out of a total of 1195 different peptides and 180 different lipids that were used for clustering, 123 proteins and 137 lipids showed significant differences between clusters. In GoDARTS, out of a total of 1195 different peptides and 199 different lipids that were used for clustering, 130 proteins and 149 lipids showed significant differences between clusters. 50 significant common peptides and 109 significant common lipids are shared between the DCS and GoDARTS cohorts (Figure 2b).

A hierarchical clustering of the relative concentration patterns of the cluster-associated features is shown as a heatmap in DCS and GoDARTS (Figure 2b; Supplemental Figure 5). In DCS, a number of discriminative omics features were observed when comparing clusters. Phosphatidylcholine, Triacylglycerol, Diacylglycerol and Ceramide were relatively higher in cluster 1; Phosphatidylcholine O and Sphingomyelin were relatively higher in cluster 2. These pronounced expression patterns were also apparent in GoDARTS. For proteins, the difference in the molecular profiles between cluster 1 and 2 can be observed in both cohorts, but the differences were more modest compared to lipids.

### GWAS analysis reveals putative associations between type 2 diabetes molecular profiles and genetic factors

Finally, in order to investigate possible genetic differences between molecular clusters, we analysed genetic loci previously associated with type 2 diabetes (17). Multi-omics clusters in each cohort were compared with type 2 diabetes SNPs using logistic regression correcting for age, BMI and gender. We did not detect any significant associations of the type 2 diabetes variants with our clusters in either cohort.

Since we had identified around 110 proteins and 130 lipids that showed altered levels between clusters, we then tested the association between the altered molecular profiles and type 2 diabetes variants. The SNP and molecular profile association was assessed in a linear regression model with SNP dosages, age, BMI and gender as independent variables. In DCS, a novel protective type 2 diabetes variant (17), rs146886108, showed putative association with several molecules such as heat shock protein 90 beta (*p* =1.08e-13) and glucose 6 phosphate isomerase (*p* = 2.04e-15). A consistent association result between rs505922 and E-selection can also be observed in both DCS (*p* = 1.23e-15) and GoDARTS (*p* = 1.40e-16) cohorts.

### Multi-Omics clusters show similarity to known clinical subgroups

We next compared the multi-omics clusters with clusters based on clinical measurements (3). Similar to a previous study (2), these clusters can be classified as severe insulin-deficient diabetes (SIDD); severe insulin-resistant diabetes (SIRD); mild obesity-related diabetes (MOD); mild diabetes (MD) and mild diabetes with high HDL (MDH). The overlap of clinical clusters with molecular clusters is shown in figure 3a. In DCS, multi-omics cluster 1 showed a significant overlap with clinical cluster SIRD and MOD. On the other hand, multi-omics cluster 2 showed a significant overlap with clinical cluster MDH. In GoDARTS, a similar trend was observed, as multi-omics cluster 1 showed a larger overlap with SIRD and MOD, and multi-omics cluster 2 showed an overlap with MDH (figure 3a).

Box plots were used to further visualize the differences between multi-omics and clinical clusters (figure 3b, Supplemental figure 4). In DCS, we observed that multi-omics cluster 1, SIRD and MOD showed similar levels of C-peptide, HOMA-2B, HOMA-2IR, and triglycerides levels. On the other hand, multi-omics cluster 2 and MDH both showed a low C-peptide and a high HDL cholesterol level (Supplemental figure 4a). A similar pattern was observed in GoDARTS (Supplemental figure 4b).

In summary, we conclude that multi-omics cluster 1 shares similar clinical features to SIRD and MOD clinical subgroups whereas multi-omics cluster 2 shows higher similarity to the MDH clinical subgroup.

## Discussion

Using an unsupervised, bottom-up approach, we were able to cluster individuals with diabetes from two large European cohorts into two robust clusters based solely on plasma-measured multi-omics data. Clinical features such as age and BMI were excluded as confounding variables driving SNF clustering as they were not significantly different between cluster 1 and cluster 2. Moreover, using six clinical features to replicate the multi-omics clustering results was unsuccessful for both cohorts. These findings suggest that patient stratification using molecular features can reveal novel insights that are complementary to clinical data.

In both the DCS and GoDARTS cohorts, patients assigned to multi-omics clusters 1 and 2 showed conserved differences in disease severity markers as measured by HOMA2-B, -IR, C-peptide and triglyceride levels, which is known to be associated with insulin resistance (20). Individuals with type 2 diabetes are at risk of many complications such as kidney disease and cardiovascular disease. It has been demonstrated that insulin resistance is associated with hallmarks of kidney damage such as glomerular hypertension, hyperfiltration and proteinuria (21, 22). We show evidence from Cox proportional hazards modelling that patients in different clusters have slower disease progression rates in both cohorts. Therefore, these multi-omics clusters could potentially define molecular signatures related to disease severity, progression and complications risk which could be relevant for more precision-based therapy of type 2 diabetes.

We tested the associations of type 2 diabetes genetic loci with multi-omics clusters but found no clear evidence to suggest that the multi-omics clusters were associated with specific type 2 diabetes genetic variants in either cohort. This suggests that known type 2 diabetes genes are not drivers of molecular differences between clusters. A similar clustering result can be observed in both cohorts and cluster 1 and cluster 2 show similar HbA1c levels suggesting that our clusters are stable and at least partially mechanistically distinct. It would be intriguing to see whether similar results can be obtained from both newly diagnosed patients and long-term diabetes patients could be performed in the future to support this view.

In both cohorts, we have identified ∼100 proteins and ∼130 lipids that showed an altered expression pattern between multi-omics cluster 1 and cluster 2. Currently, we cannot fully identify which molecules likely to be the driver molecules, nor we can say these molecules play an important role in type 2 diabetes development. Nevertheless, we assessed the association between these molecules and the type 2 diabetes variants. A novel type 2 diabetes SNP, rs146886108, a missense variant that encodes p.Arg187Gln in ANKH, which was first identified by Mahajan et a. (17), and showed a strong correlation with several measured proteins, including heat shock protein 90 beta, glucose 6 phosphate isomerase. The UK Biobank Mendelian trait GWAS study also showed a strong association between rs146886108 and random glucose. The fact that we found a strong association between rs146886108 and glucose 6 phosphate isomerase could potentially identify a mechanism behind the action of rs146886108 on glucose regulation. Further investigations may help to evaluate the causal roles of these type 2 diabetes variants, with their associated molecules and possible mechanisms.

Using five clinical measurements, age, BMI, HbA1c, c-peptide and HDL, Slieker and Donnelly et al. (3) have shown that people with type 2 diabetes could be grouped into five distinct clusters: insulin-deficient (SIDD), insulin-resistant (SIRD), an obesity cluster (MOD), mild diabetes (MD) and MD with a high HDL (MDH). Comparing with these clinical clusters, we were able to demonstrate that the multi-omics clusters significantly overlap with the clinically defined clusters. Multi-omics cluster 1, characterized by a relatively high C-peptide and triglycerides level, showed a significant overlap with SIRD and MOD. Multi-omics cluster 2, a cluster with a relatively high HDL and low C-peptide value, showed a significant overlap with MDH. These results further underline the complementarity of the multi-omics and clinical clustering approaches and suggests that multi-omics signatures can in theory be linked to clinical measurements and outcomes to explore molecular mechanisms linked to the underlying heterogeneity of disease.

The present approach offers the prospect of multi-omics personalized medicine, in which an extended set of molecular characteristics is used to assess disease development and progression. Through multi-omics health management, each patient might be positioned with respect to the underlying dysregulated pathways, with therapies selected accordingly. Whether this approach will indeed provide better outcomes than current practice, based on clinical features and more limited blood biochemistry, will need to be assessed in the future.

## Limitations of study

From the patient network (Figure 1) it is clear that the multi-omics clusters are not completely distinct. The two clusters therefore represent different molecular signatures, where a patient shows more similarity to one cluster compared to the other on a continuous scale. Nevertheless, we do not recommend using such clusters to assign patients, but rather the use of the clustering results to explore the underlying molecular pathology and as an alternative route to identifying novel biomarkers for risk of type 2 diabetes and its complications.

In both DCS and GoDARTS, patients were not necessarily recruited after diagnosis, and this heterogeneity in the disease duration could potentially have an effect on the blood measurements obtained. Moreover, as we mentioned above, in DCS, blood samples are taken in a fasted state, in contrast to GoDARTS, where blood measurements are done in a non-fasted state. However, it has been demonstrated that a normal food intake only has slight effects on cholesterol levels, and lipid profiles at most change minimally in response to normal food intake in general populations (23, 24).

One of the common questions regarding patients’ clustering results is whether patients move between clusters over time. In a recent 5-year follow-up study (25), 367 diabetes patients were assigned based on the nearest centroid approaches at both baseline and 5-year follow-up. 23% of patients switched their cluster allocations after 5 years. This can be expected as the variables that were used for clustering such as age, BMI and HOMA2 can change over time, following the time increase, lifestyle choice changes and disease progress. For our study, roughly 1375 plasma molecules were used for clustering. Some molecule concentrations such as triacylglycerol and diacylglycerol can change following lifestyle changes or medical treatment. Other “housekeeping” molecules are more likely to remain constant over time. The stability of molecular clustering results needs to be assessed in further prospective studies.

We cannot at this stage assert that the different clusters represent different aetiologies of type 2 diabetes, nor that the clustering method represents an optimal classification for type 2 diabetes subgroups. The multi-omics clusters in our study were derived primarily from patients from Europe. Consequently, the applicability of these results to other ethnic groups needs to be assessed.

## Supporting information

Conflict of interest disclosure

STROBE checklist

## Data Availability

All data produced in the present study are available upon reasonable request to the authors.

## Acknowledgments

The authors acknowledge the support of the Health Informatics Centre, University of Dundee, for managing and supplying the anonymized data.

## Contribution Statement

S.L., M.I. and G.A.R. designed the research. S.L. wrote the manuscript. S.L. performed all data analysis with technical support from I.D.. I.D., D.K., and M.I. set up a federated node system for data analysis. R.C.S., L.A.D., M.K.H., J.W.J.B., L.M’t.H., F..M, P.J.M.E. and E.R.P. were involved in generating the peptidomics data, generating the clinical cluster assignment results, managing and handling the data in databases. M.JG., C.K. and K.S. generated the Lipotype data. The research was designed partly based on the pre-work results by C.H.F. All authors read and approved the manuscript and contributed according to the ICMJE criteria (www.icmje.org/). G.A.R. and M.I. co-supervised the work. M.I. is the guarantor of this work and, as such, had full access to all the data in the study and takes responsibility for the integrity of the data and the accuracy of the data analysis.

**Table 1.** Genetic associations with altered molecular profiles between multiomics clusters reaching significance. The correlations between molecules and SNPs were estimated using linear regression models with age, BMI and gender adjustments. The commonly accepted threshold P ≤ 5 × 10(-8) for genome-wide association studies is applied in this present study.

| DCS |  |  |  |  |  |  |
| --- | --- | --- | --- | --- | --- | --- |
| Locus | rs ID | Chr | Pos | Alleles | association molecule | P-value (adjusted by BMI, age and sex) |
| ANKH | rs146886108 | 5 | 14,751,305 | C/T | Integrin alpha I beta 1 complex | 5.88E-10 |
| ANKH | rs146886108 | 5 | 14,751,305 | C/T | Heat shock protein HSP 90 alpha beta | 1.13E-21 |
| ANKH | rs146886108 | 5 | 14,751,305 | C/T | Heat shock protein HSP 90 beta | 1.08E-13 |
| ANKH | rs146886108 | 5 | 14,751,305 | C/T | Glucose 6 phosphate isomerase | 2.04E-15 |
| ANKH | rs146886108 | 5 | 14,751,305 | C/T | Alcohol dehydrogenase NADP | 4.04E-32 |
| ABO | rs505922 | 9 | 133,273,813 | C/T | E selectin | 1.23E-15 |
| ABO | rs505922 | 9 | 133,273,813 | C/T | CD209 antigen | 2.09E-14 |
| **GoDARTS** |  |  |  |  |  |  |
| **Locus** | **rs ID** | **Chr** | **Pos** | **Alleles** | **association molecule** | **P-value (adjusted by BMI, age and sex)** |
| ABO | rs505922 | 9 | 133,273,813 | C/T | E selectin | 1.40E-16 |

