## Supplemental Figures for "Novel subgroups of type 2 diabetes based on multi-Omics profiling: an IMI-RHAPSODY Study"

Supplemental Figure 1

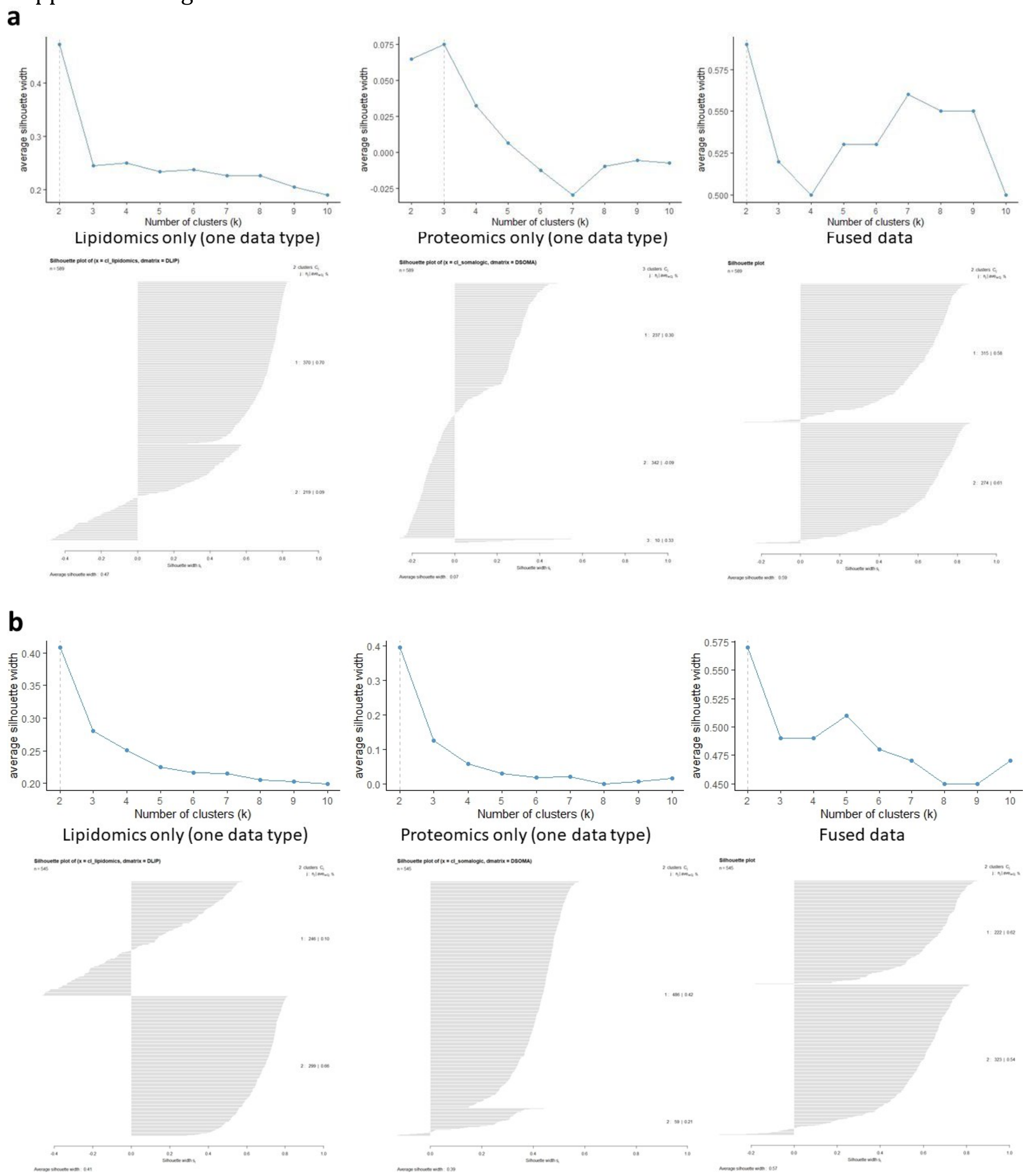


**Supplemental Figure 1. Silhouette results for each of the omics data independently and the integrated multi-omics data.** The silhouette method was performed on both cohorts with the number of clusters ($k$) ranging from 2 to 10. The silhouette method is used to evaluate how well each patient lies within their assigned cluster compared to its neighbour cluster. A high silhouette width value indicates the samples are well matched to their clusters. The silhouette plots of the optimal assignment results (the highest average silhouette width) for each data type were also shown to reflect the silhouette width of each individual. **(a)** The silhouette method was performed on the DCS cohort consisting of 589 T2D patients. (**b**) The silhouette method was performed on the GoDARTS cohort consisting of 545 T2D patients.

Supplemental Figure 2

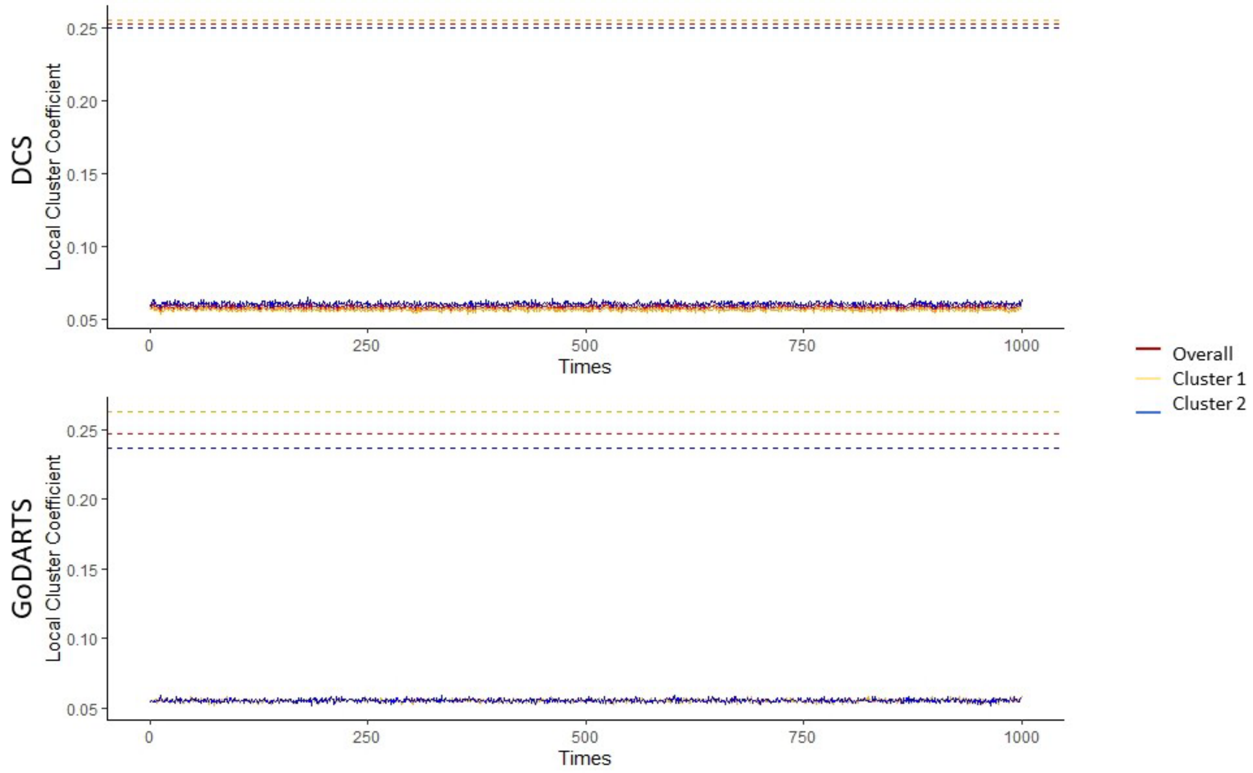


**Supplemental Figure 2. Multi-omics clustering results were validated by bootstrapping (n=1000 iterations) test.** The simulated models were iterated 1000 times to record the frequency of achieving a model with an equal or greater mean local cluster coefficient than the study model. For the simulated data, the same parameters were used and the number of clusters is set to correspond to the number of clusters in the study model. The dash lines indicate the local coefficient of the study models.

Supplemental Figure 3

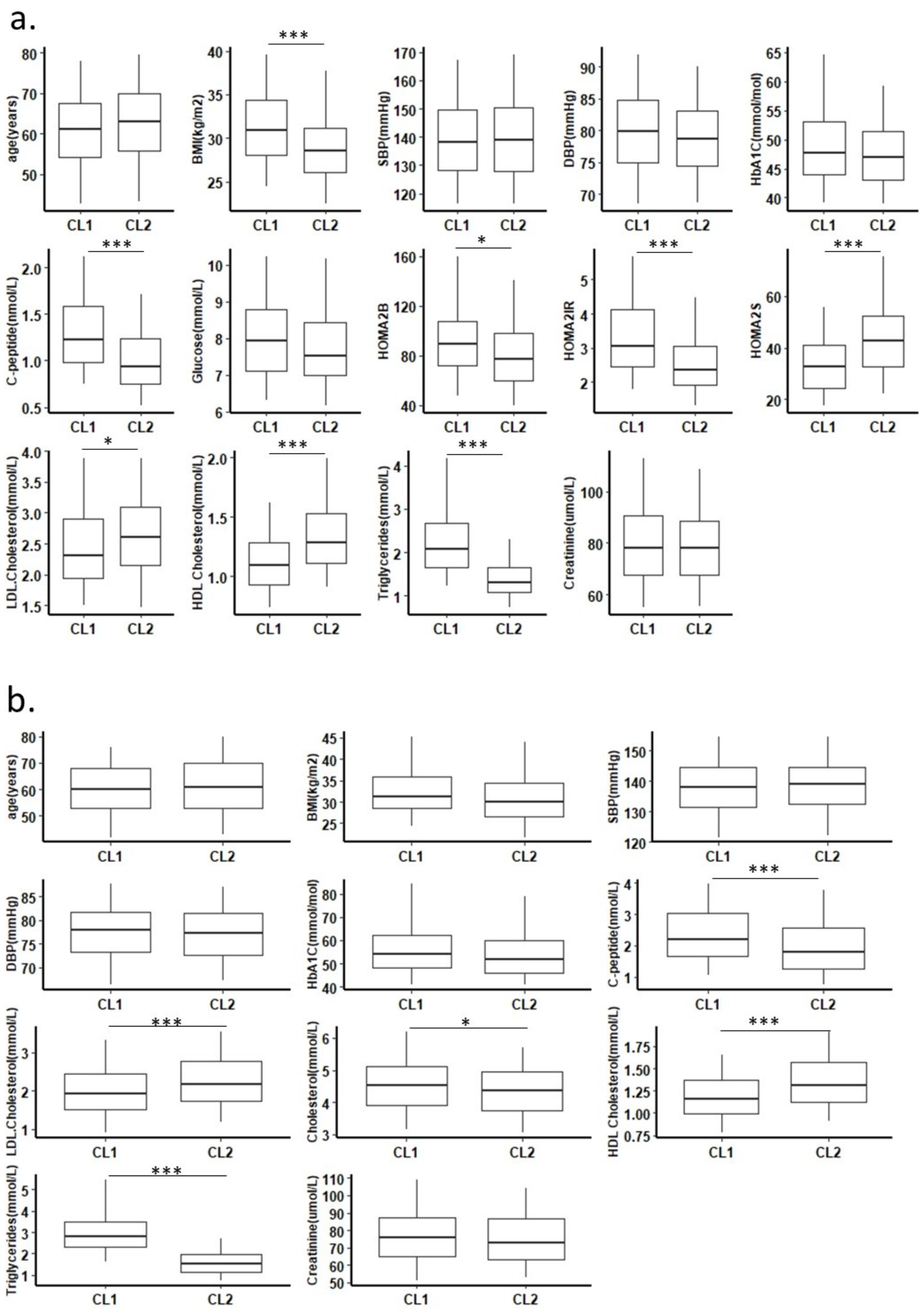

**ESM Figure 3. Clinical features of multi-omics clusters in both DCS and GoDARTS cohorts.** Distributions of clinical measurements at baseline in DCS (**a**) and GoDARTS (**b**) for multi-omics cluster #1 and cluster #2. SBP=systolic blood pressure. DBP=diastolic blood pressure. HbA1C=Hemoglobin A1C. GAD1=Glutamic Acid Decarboxylase 1. SIRD= severe insulin-resistant diabetes. CL1=multi-omics cluster #1. CL2=multi-omics cluster #2. *:P-value<=0.05. **:P-value<=0.01. ***:P- value<=0.001.

Supplemental Figure 4

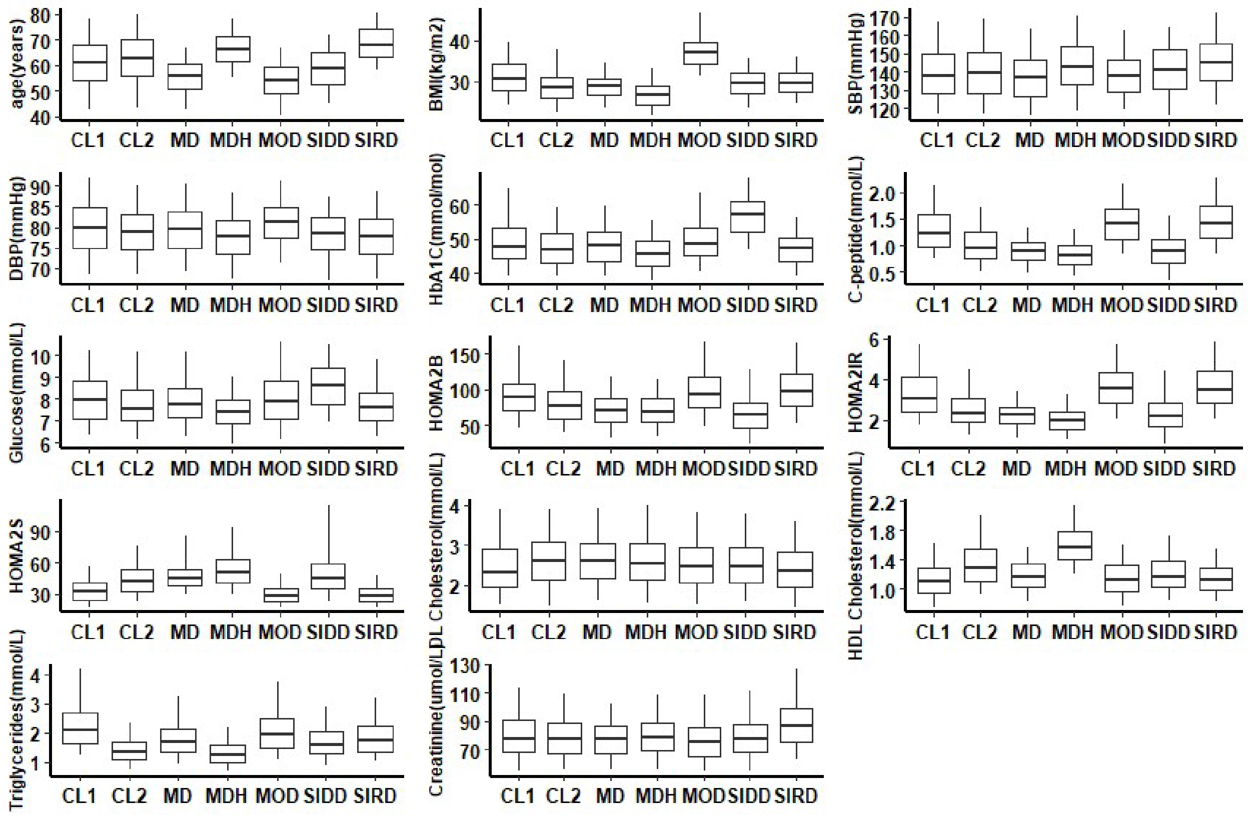

b.

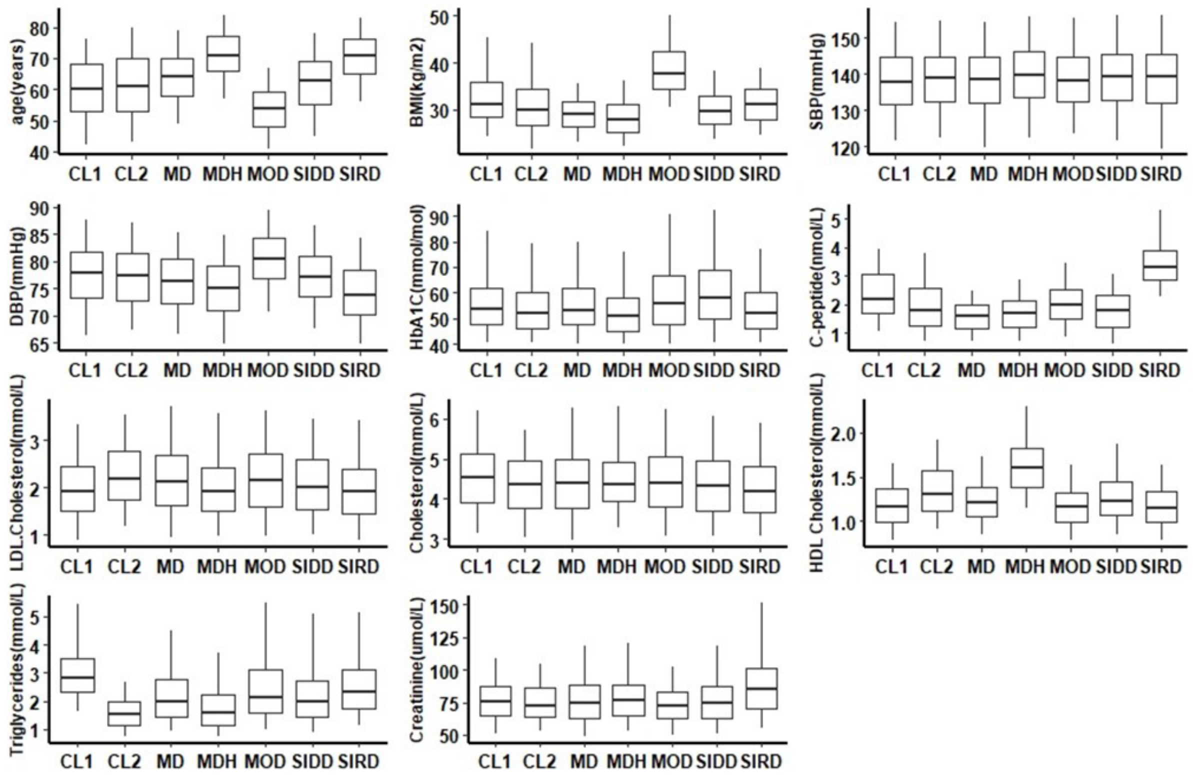


**Supplemental Figure 4. Clinical features of multi-omics clusters and the clinical clusters in both DCS and GoDARTS cohorts.** Distributions of clinical measurements at baseline in DCS (**a**) and GoDARTS (**b**). CL1= multi-omics cluster #1. CL2= multi- omics cluster #2. MD= mild diabetes. MDH=mild diabetes with high HDL. MOD= mild obesity-related diabetes. SIDD= severely insulin-deficient diabetes. SIRD= severely insulin-resistant diabetes.

Supplemental figure 5

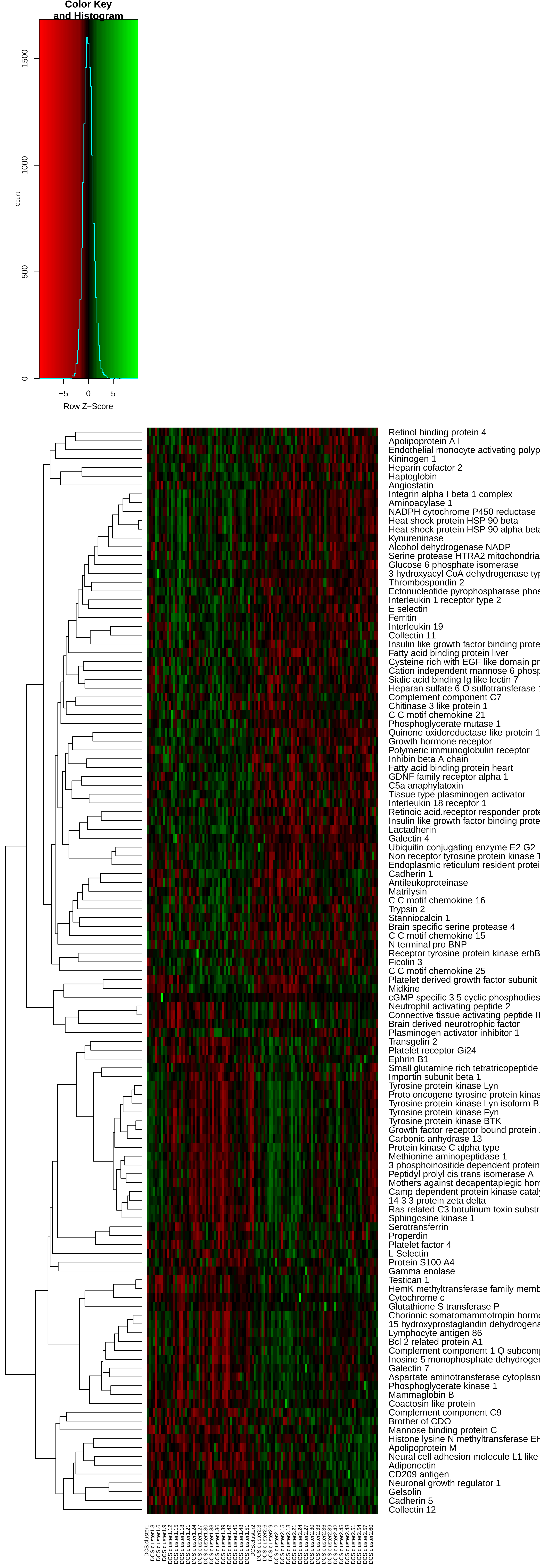

Color Key
and Histogram
Count
1500
1000
500
0
−5
0
5
Row Z−Score
Retinol binding protein 4
Apolipoprotein A I
Endothelial monocyte activating polyp
Kininogen 1
Heparin cofactor 2
Haptoglobin
Angiostatin
Integrin alpha I beta 1 complex
Aminoacylase 1
NADPH cytochrome P450 reductase
Heat shock protein HSP 90 beta
Heat shock protein HSP 90 alpha beta
Kynureninase
Alcohol dehydrogenase NADP
Serine protease HTRA2 mitochondria
Glucose 6 phosphate isomerase
3 hydroxyacyl CoA dehydrogenase typ
Thrombospondin 2
Ectonucleotide pyrophosphatase phos
Interleukin 1 receptor type 2
E selectin
Ferritin
Interleukin 19
Collectin 11
Insulin like growth factor binding prote
Fatty acid binding protein liver
Cysteine rich with EGF like domain pr
Cation independent mannose 6 phosp
Sialic acid binding Ig like lectin 7
Heparan sulfate 6 O sulfotransferase 1
Complement component C7
Chitinase 3 like protein 1
C C motif chemokine 21
Phosphoglycerate mutase 1
Quinone oxidoreductase like protein 1
Growth hormone receptor
Polymeric immunoglobulin receptor
Inhibin beta A chain
Fatty acid binding protein heart
GDNF family receptor alpha 1
C5a anaphylatoxin
Tissue type plasminogen activator
Interleukin 18 receptor 1
Retinoic acid.receptor responder prote
Insulin like growth factor binding prote
Lactadherin
Galectin 4
Ubiquitin conjugating enzyme E2 G2
Non receptor tyrosine protein kinase T
Endoplasmic reticulum resident protei
Cadherin 1
Antileukoproteinase
Matrilysin
C C motif chemokine 16
Trypsin 2
Stanniocalcin 1
Brain specific serine protease 4
C C motif chemokine 15
N terminal pro BNP
Receptor tyrosine protein kinase erbB
Ficolin 3
C C motif chemokine 25
Platelet derived growth factor subunit
Midkine
cGMP specific 3 5 cyclic phosphodies
Neutrophil activating peptide 2
Connective tissue activating peptide II
Brain derived neurotrophic factor
Plasminogen activator inhibitor 1
Transgelin 2
Platelet receptor Gi24
Ephrin B1
Small glutamine rich tetratricopeptide
Importin subunit beta 1
Tyrosine protein kinase Lyn
Proto oncogene tyrosine protein kinas
Tyrosine protein kinase Lyn isoform B
Tyrosine protein kinase Fyn
Tyrosine protein kinase BTK
Growth factor receptor bound protein
Carbonic anhydrase 13
Protein kinase C alpha type
Methionine aminopeptidase 1
3 phosphoinositide dependent protein
Peptidyl prolyl cis trans isomerase A
Mothers against decapentaplegic hom
Camp dependent protein kinase catal
14 3 3 protein zeta delta
Ras related C3 botulinum toxin substr
Sphingosine kinase 1
Serotransferrin
Properdin
Platelet factor 4
L Selectin
Protein S100 A4
Gamma enolase
Testican 1
HemK methyltransferase family memb
Cytochrome c
Glutathione S transferase P
Chorionic somatomammotropin hormo
15 hydroxyprostaglandin dehydrogena
Lymphocyte antigen 86
Bcl 2 related protein A1
Complement component 1 Q subcomp
Inosine 5 monophosphate dehydroger
Galectin 7
Aspartate aminotransferase cytoplasn
Phosphoglycerate kinase 1
Mammaglobin B
Coactosin like protein
Complement component C9
Brother of CDO
Mannose binding protein C
Histone lysine N methyltransferase EH
Apolipoprotein M
Neural cell adhesion molecule L1 like
Adiponectin
CD209 antigen
Neuronal growth regulator 1
Gelsolin
Cadherin 5
Collectin 12

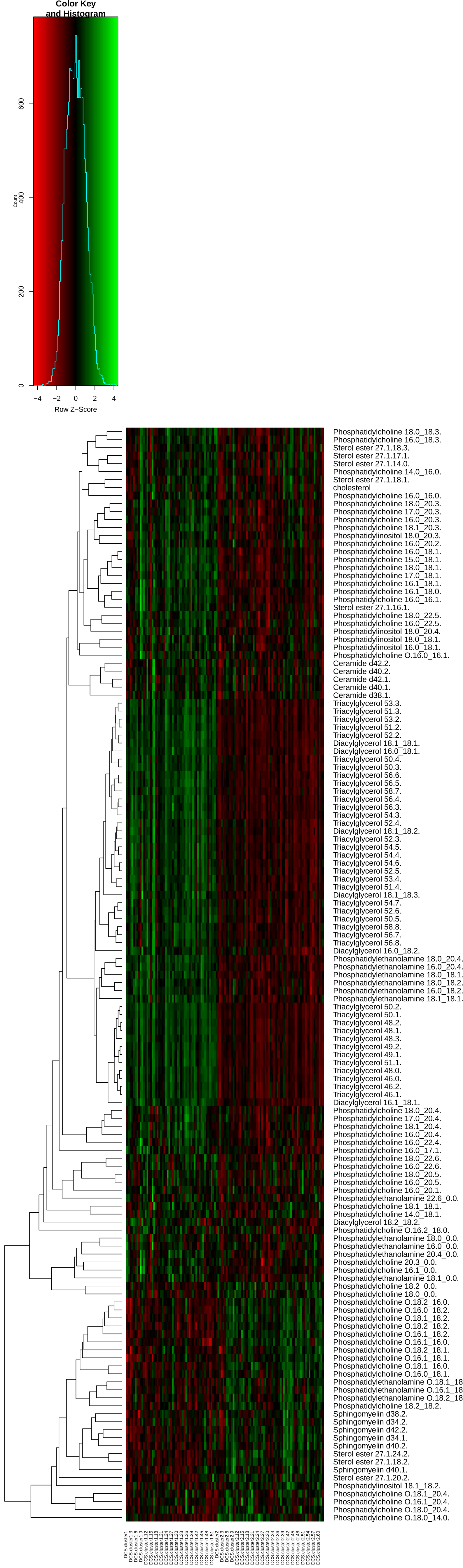
Color Key
and Histogram
Count
Row Z−Score
Phosphatidylcholine 18.0_18.3.
Phosphatidylcholine 16.0_18.3.
Sterol ester 27.1.18.3.
Sterol ester 27.1.17.1.
Sterol ester 27.1.14.0.
Phosphatidylcholine 14.0_16.0.
Sterol ester 27.1.18.1.
cholesterol
Phosphatidylcholine 16.0_16.0.
Phosphatidylcholine 18.0_20.3.
Phosphatidylcholine 17.0_20.3.
Phosphatidylcholine 16.0_20.3.
Phosphatidylcholine 18.1_20.3.
Phosphatidylinositol 18.0_20.3.
Phosphatidylcholine 16.0_20.2.
Phosphatidylcholine 16.0_18.1.
Phosphatidylcholine 15.0_18.1.
Phosphatidylcholine 18.0_18.1.
Phosphatidylcholine 17.0_18.1.
Phosphatidylcholine 16.1_18.1.
Phosphatidylcholine 16.1_18.0.
Phosphatidylcholine 16.0_16.1.
Sterol ester 27.1.16.1.
Phosphatidylcholine 18.0_22.5.
Phosphatidylcholine 16.0_22.5.
Phosphatidylinositol 18.0_20.4.
Phosphatidylinositol 18.0_18.1.
Phosphatidylinositol 16.0_18.1.
Phosphatidylcholine O.16.0_16.1.
Ceramide d42.2.
Ceramide d40.2.
Ceramide d42.1.
Ceramide d40.1.
Ceramide d38.1.
Triacylglycerol 53.3.
Triacylglycerol 51.3.
Triacylglycerol 53.2.
Triacylglycerol 51.2.
Triacylglycerol 52.2.
Diacylglycerol 18.1_18.1.
Diacylglycerol 16.0_18.1.
Triacylglycerol 50.4.
Triacylglycerol 50.3.
Triacylglycerol 56.6.
Triacylglycerol 56.5.
Triacylglycerol 58.7.
Triacylglycerol 56.4.
Triacylglycerol 56.3.
Triacylglycerol 54.3.
Triacylglycerol 52.4.
Diacylglycerol 18.1_18.2.
Triacylglycerol 52.3.
Triacylglycerol 54.5.
Triacylglycerol 54.4.
Triacylglycerol 54.6.
Triacylglycerol 52.5.
Triacylglycerol 53.4.
Triacylglycerol 51.4.
Diacylglycerol 18.1_18.3.
Triacylglycerol 54.7.
Triacylglycerol 52.6.
Triacylglycerol 50.5.
Triacylglycerol 58.8.
Triacylglycerol 56.7.
Triacylglycerol 56.8.
Diacylglycerol 16.0_18.2.
Phosphatidylethanolamine 18.0_20.4.
Phosphatidylethanolamine 16.0_20.4.
Phosphatidylethanolamine 18.0_18.1.
Phosphatidylethanolamine 18.0_18.2.
Phosphatidylethanolamine 16.0_18.2.
Phosphatidylethanolamine 18.1_18.1.
Triacylglycerol 50.2.
Triacylglycerol 50.1.
Triacylglycerol 48.2.
Triacylglycerol 48.1.
Triacylglycerol 48.3.
Triacylglycerol 49.2.
Triacylglycerol 49.1.
Triacylglycerol 51.1.
Triacylglycerol 48.0.
Triacylglycerol 46.0.
Triacylglycerol 46.2.
Triacylglycerol 46.1.
Diacylglycerol 16.1_18.1.
Phosphatidylcholine 18.0_20.4.
Phosphatidylcholine 17.0_20.4.
Phosphatidylcholine 18.1_20.4.
Phosphatidylcholine 16.0_20.4.
Phosphatidylcholine 16.0_22.4.
Phosphatidylcholine 16.0_17.1.
Phosphatidylcholine 18.0_22.6.
Phosphatidylcholine 16.0_22.6.
Phosphatidylcholine 18.0_20.5.
Phosphatidylcholine 16.0_20.5.
Phosphatidylcholine 16.0_20.1.
Phosphatidylethanolamine 22.6_0.0.
Phosphatidylcholine 18.1_18.1.
Phosphatidylcholine 14.0_18.1.
Diacylglycerol 18.2_18.2.
Phosphatidylcholine O.16.2_18.0.
Phosphatidylethanolamine 18.0_0.0.
Phosphatidylethanolamine 16.0_0.0.
Phosphatidylethanolamine 20.4_0.0.
Phosphatidylcholine 20.3_0.0.
Phosphatidylcholine 16.1_0.0.
Phosphatidylethanolamine 18.1_0.0.
Phosphatidylcholine 18.2_0.0.
Phosphatidylcholine 18.0_0.0.
Phosphatidylcholine O.18.2_16.0.
Phosphatidylcholine O.16.0_18.2.
Phosphatidylcholine O.18.1_18.2.
Phosphatidylcholine O.18.2_18.2.
Phosphatidylcholine O.16.1_18.2.
Phosphatidylcholine O.16.1_16.0.
Phosphatidylcholine O.18.2_18.1.
Phosphatidylcholine O.16.1_18.1.
Phosphatidylcholine O.18.1_16.0.
Phosphatidylcholine O.16.0_18.1.
Phosphatidylethanolamine O.18.1_18
Phosphatidylethanolamine O.16.1_18
Phosphatidylethanolamine O.18.2_18
Phosphatidylcholine 18.2_18.2.
Sphingomyelin d38.2.
Sphingomyelin d34.2.
Sphingomyelin d42.2.
Sphingomyelin d34.1.
Sphingomyelin d40.2.
Sterol ester 27.1.24.2.
Sterol ester 27.1.18.2.
Sphingomyelin d40.1.
Sterol ester 27.1.20.2.
Phosphatidylinositol 18.1_18.2.
Phosphatidylcholine O.18.1_20.4.
Phosphatidylcholine O.16.1_20.4.
Phosphatidylcholine O.18.0_20.4.
Phosphatidylcholine O.18.0_14.0.

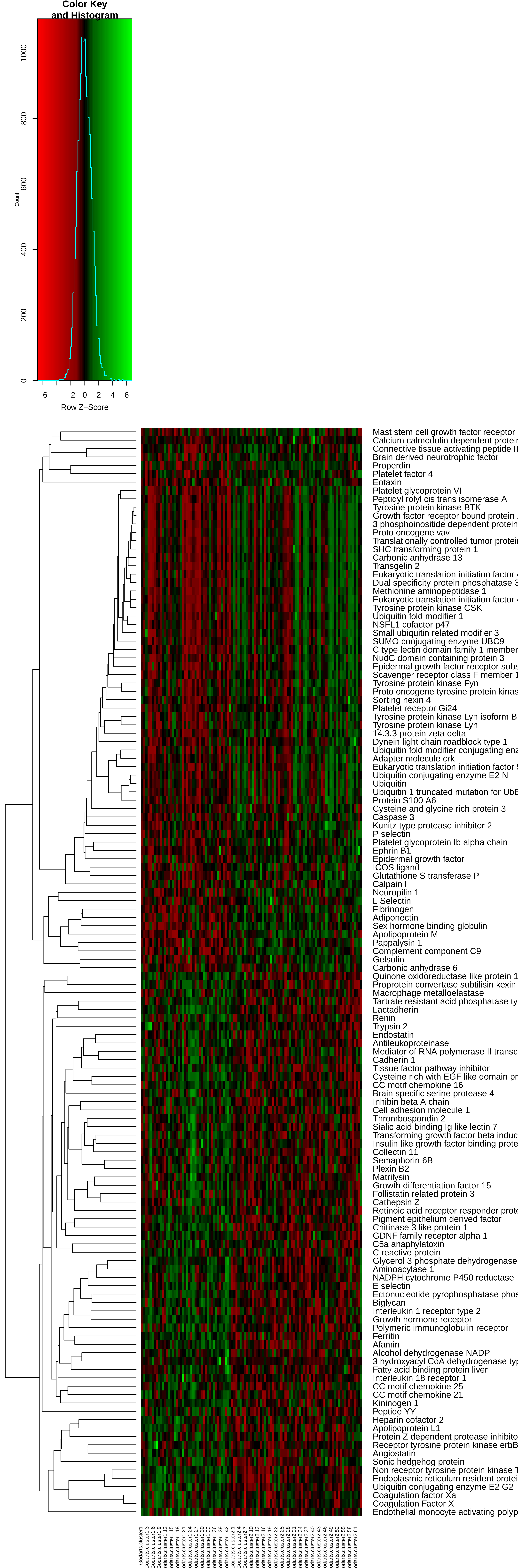

Color Key
and Histogram
Count
0
200
400
600
800
1000
−6
−2
0
2
4
6
Row Z−Score
Mast stem cell growth factor receptor
Calcium calmodulin dependent protein
Connective tissue activating peptide II
Brain derived neurotrophic factor
Properdin
Platelet factor 4
Eotaxin
Platelet glycoprotein VI
Peptidyl rolyl cis trans isomerase A
Tyrosine protein kinase BTK
Growth factor receptor bound protein
3 phosphoinositide dependent protein
Proto oncogene vav
Translationally controlled tumor protein
SHC transforming protein 1
Carbonic anhydrase 13
Transgelin 2
Eukaryotic translation initiation factor
Dual specificity protein phosphatase
Methionine aminopeptidase 1
Eukaryotic translation initiation factor
Tyrosine protein kinase CSK
Ubiquitin fold modifier 1
NSFL1 cofactor p47
Small ubiquitin related modifier 3
SUMO conjugating enzyme UBC9
C type lectin domain family 1 member
NudC domain containing protein 3
Epidermal growth factor receptor subs
Scavenger receptor class F member 1
Tyrosine protein kinase Fyn
Proto oncogene tyrosine protein kinas
Sorting nexin 4
Platelet receptor Gi24
Tyrosine protein kinase Lyn isoform B
Tyrosine protein kinase Lyn
14.3.3 protein zeta delta
Dynein light chain roadblock type 1
Ubiquitin fold modifier conjugating enz
Adapter molecule crk
Eukaryotic translation initiation factor
Ubiquitin conjugating enzyme E2 N
Ubiquitin
Ubiquitin 1 truncated mutation for UbE
Protein S100 A6
Cysteine and glycine rich protein 3
Caspase 3
Kunitz type protease inhibitor 2
P selectin
Platelet glycoprotein Ib alpha chain
Ephrin B1
Epidermal growth factor
ICOS ligand
Glutathione S transferase P
Calpain I
Neuropilin 1
L Selectin
Fibrinogen
Adiponectin
Sex hormone binding globulin
Apolipoprotein M
Pappalysin 1
Complement component C9
Gelsolin
Carbonic anhydrase 6
Quinone oxidoreductase like protein 1
Proprotein convertase subtilisin kexin
Macrophage metalloelastase
Tartrate resistant acid phosphatase ty
Lactadherin
Renin
Trypsin 2
Endostatin
Antileukoproteinase
Mediator of RNA polymerase II transc
Cadherin 1
Tissue factor pathway inhibitor
Cysteine rich with EGF like domain pr
CC motif chemokine 16
Brain specific serine protease 4
Inhibin beta A chain
Cell adhesion molecule 1
Thrombospondin 2
Sialic acid binding Ig like lectin 7
Transforming growth factor beta induc
Insulin like growth factor binding prote
Collectin 11
Semaphorin 6B
Plexin B2
Matrilysin
Growth differentiation factor 15
Follistatin related protein 3
Cathepsin Z
Retinoic acid receptor responder prote
Pigment epithelium derived factor
Chitinase 3 like protein 1
GDNF family receptor alpha 1
C5a anaphylatoxin
C reactive protein
Glycerol 3 phosphate dehydrogenase
Aminoacylase 1
NADPH cytochrome P450 reductase
E selectin
Ectonucleotide pyrophosphatase phos
Biglycan
Interleukin 1 receptor type 2
Growth hormone receptor
Polymeric immunoglobulin receptor
Ferritin
Afamin
Alcohol dehydrogenase NADP
3 hydroxyacyl CoA dehydrogenase typ
Fatty acid binding protein liver
Interleukin 18 receptor 1
CC motif chemokine 25
CC motif chemokine 21
Kininogen 1
Peptide YY
Heparin cofactor 2
Apolipoprotein L1
Protein Z dependent protease inhibito
Receptor tyrosine protein kinase erbB
Angiostatin
Sonic hedgehog protein
Non receptor tyrosine protein kinase T
Endoplasmic reticulum resident protei
Ubiquitin conjugating enzyme E2 G2
Coagulation factor Xa
Coagulation Factor X
Endothelial monocyte activating polyp

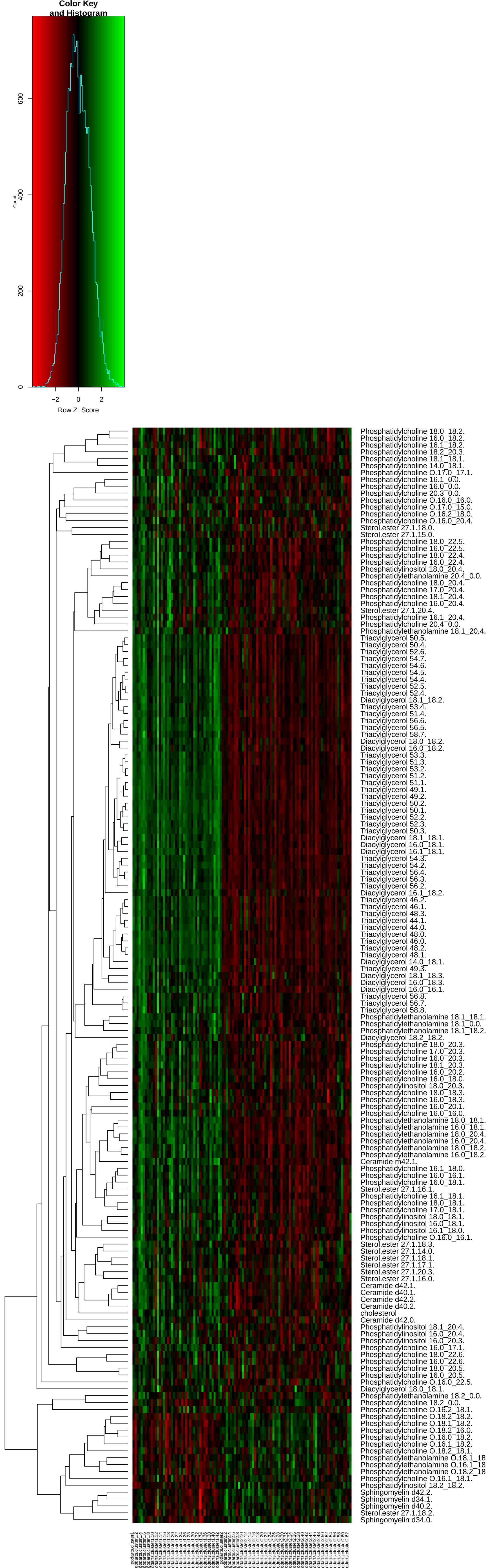

Color Key
and Histogram
Count
600
400
200
0
−2
0
2
Row Z−Score
Phosphatidylcholine 18.0_18.2.
Phosphatidylcholine 16.0_18.2.
Phosphatidylcholine 16.1_18.2.
Phosphatidylcholine 18.2_20.3.
Phosphatidylcholine 18.1_18.1.
Phosphatidylcholine 14.0_18.1.
Phosphatidylcholine O.17.0_17.1.
Phosphatidylcholine 16.1_0.0.
Phosphatidylcholine 16.0_0.0.
Phosphatidylcholine 20.3_0.0.
Phosphatidylcholine O.16.0_16.0.
Phosphatidylcholine O.17.0_15.0.
Phosphatidylcholine O.16.2_18.0.
Phosphatidylcholine O.16.0_20.4.
Sterol.ester 27.1.18.0.
Sterol.ester 27.1.15.0.
Phosphatidylcholine 18.0_22.5.
Phosphatidylcholine 16.0_22.5.
Phosphatidylcholine 18.0_22.4.
Phosphatidylcholine 16.0_22.4.
Phosphatidylinositol 18.0_20.4.
Phosphatidylethanolamine 20.4_0.0.
Phosphatidylcholine 18.0_20.4.
Phosphatidylcholine 17.0_20.4.
Phosphatidylcholine 18.1_20.4.
Phosphatidylcholine 16.0_20.4.
Sterol.ester 27.1.20.4.
Phosphatidylcholine 16.1_20.4.
Phosphatidylcholine 20.4_0.0.
Phosphatidylethanolamine 18.1_20.4.
Triacylglycerol 50.5.
Triacylglycerol 50.4.
Triacylglycerol 52.6.
Triacylglycerol 54.7.
Triacylglycerol 54.6.
Triacylglycerol 54.5.
Triacylglycerol 54.4.
Triacylglycerol 52.5.
Triacylglycerol 52.4.
Diacylglycerol 18.1_18.2.
Triacylglycerol 53.4.
Triacylglycerol 51.4.
Triacylglycerol 56.6.
Triacylglycerol 56.5.
Triacylglycerol 58.7.
Diacylglycerol 18.0_18.2.
Diacylglycerol 16.0_18.2.
Triacylglycerol 53.3.
Triacylglycerol 51.3.
Triacylglycerol 53.2.
Triacylglycerol 51.2.
Triacylglycerol 51.1.
Triacylglycerol 49.1.
Triacylglycerol 49.2.
Triacylglycerol 50.2.
Triacylglycerol 50.1.
Triacylglycerol 52.2.
Triacylglycerol 52.3.
Triacylglycerol 50.3.
Diacylglycerol 18.1_18.1.
Diacylglycerol 16.0_18.1.
Diacylglycerol 16.1_18.1.
Triacylglycerol 54.3.
Triacylglycerol 54.2.
Triacylglycerol 56.4.
Triacylglycerol 56.3.
Triacylglycerol 56.2.
Diacylglycerol 16.1_18.2.
Triacylglycerol 46.2.
Triacylglycerol 46.1.
Triacylglycerol 48.3.
Triacylglycerol 44.1.
Triacylglycerol 44.0.
Triacylglycerol 48.0.
Triacylglycerol 46.0.
Triacylglycerol 48.2.
Triacylglycerol 48.1.
Diacylglycerol 14.0_18.1.
Triacylglycerol 49.3.
Diacylglycerol 18.1_18.3.
Diacylglycerol 16.0_18.3.
Diacylglycerol 16.0_16.1.
Triacylglycerol 56.8.
Triacylglycerol 56.7.
Triacylglycerol 58.8.
Phosphatidylethanolamine 18.1_18.1.
Phosphatidylethanolamine 18.1_0.0.
Phosphatidylethanolamine 18.1_18.2.
Diacylglycerol 18.2_18.2.
Phosphatidylcholine 18.0_20.3.
Phosphatidylcholine 17.0_20.3.
Phosphatidylcholine 16.0_20.3.
Phosphatidylcholine 18.1_20.3.
Phosphatidylcholine 16.0_20.2.
Phosphatidylcholine 16.0_18.0.
Phosphatidylinositol 18.0_20.3.
Phosphatidylcholine 18.0_18.3.
Phosphatidylcholine 16.0_18.3.
Phosphatidylcholine 16.0_20.1.
Phosphatidylcholine 16.0_16.0.
Phosphatidylethanolamine 18.0_18.1.
Phosphatidylethanolamine 16.0_18.1.
Phosphatidylethanolamine 18.0_20.4.
Phosphatidylethanolamine 16.0_20.4.
Phosphatidylethanolamine 18.0_18.2.
Phosphatidylethanolamine 16.0_18.2.
Ceramide m42.1.
Phosphatidylcholine 16.1_18.0.
Phosphatidylcholine 16.0_16.1.
Phosphatidylcholine 16.0_18.1.
Sterol.ester 27.1.16.1.
Phosphatidylcholine 16.1_18.1.
Phosphatidylcholine 18.0_18.1.
Phosphatidylcholine 17.0_18.1.
Phosphatidylinositol 18.0_18.1.
Phosphatidylinositol 16.0_18.1.
Phosphatidylinositol 16.1_18.0.
Phosphatidylcholine O.16.0_16.1.
Sterol.ester 27.1.18.3.
Sterol.ester 27.1.14.0.
Sterol.ester 27.1.18.1.
Sterol.ester 27.1.17.1.
Sterol.ester 27.1.20.3.
Sterol.ester 27.1.16.0.
Ceramide d42.1.
Ceramide d40.1.
Ceramide d42.2.
Ceramide d40.2.
cholesterol
Ceramide d42.0.
Phosphatidylinositol 18.1_20.4.
Phosphatidylinositol 16.0_20.4.
Phosphatidylinositol 16.0_20.3.
Phosphatidylcholine 16.0_17.1.
Phosphatidylcholine 18.0_22.6.
Phosphatidylcholine 16.0_22.6.
Phosphatidylcholine 18.0_20.5.
Phosphatidylcholine 16.0_20.5.
Phosphatidylcholine O.16.0_22.5.
Diacylglycerol 18.0_18.1.
Phosphatidylethanolamine 18.2_0.0.
Phosphatidylcholine 18.2_0.0.
Phosphatidylcholine O.16.2_18.1.
Phosphatidylcholine O.18.2_18.2.
Phosphatidylcholine O.18.1_18.2.
Phosphatidylcholine O.18.2_16.0.
Phosphatidylcholine O.16.0_18.2.
Phosphatidylcholine O.16.1_18.2.
Phosphatidylcholine O.18.2_18.1.
Phosphatidylethanolamine O.18.1_18
Phosphatidylethanolamine O.16.1_18
Phosphatidylethanolamine O.18.2_18
Phosphatidylcholine O.16.1_18.1.
Phosphatidylinositol 18.2_18.2.
Sphingomyelin d42.2.
Sphingomyelin d34.1.
Sphingomyelin d40.2.
Sterol.ester 27.1.18.2.
Sphingomyelin d34.0.

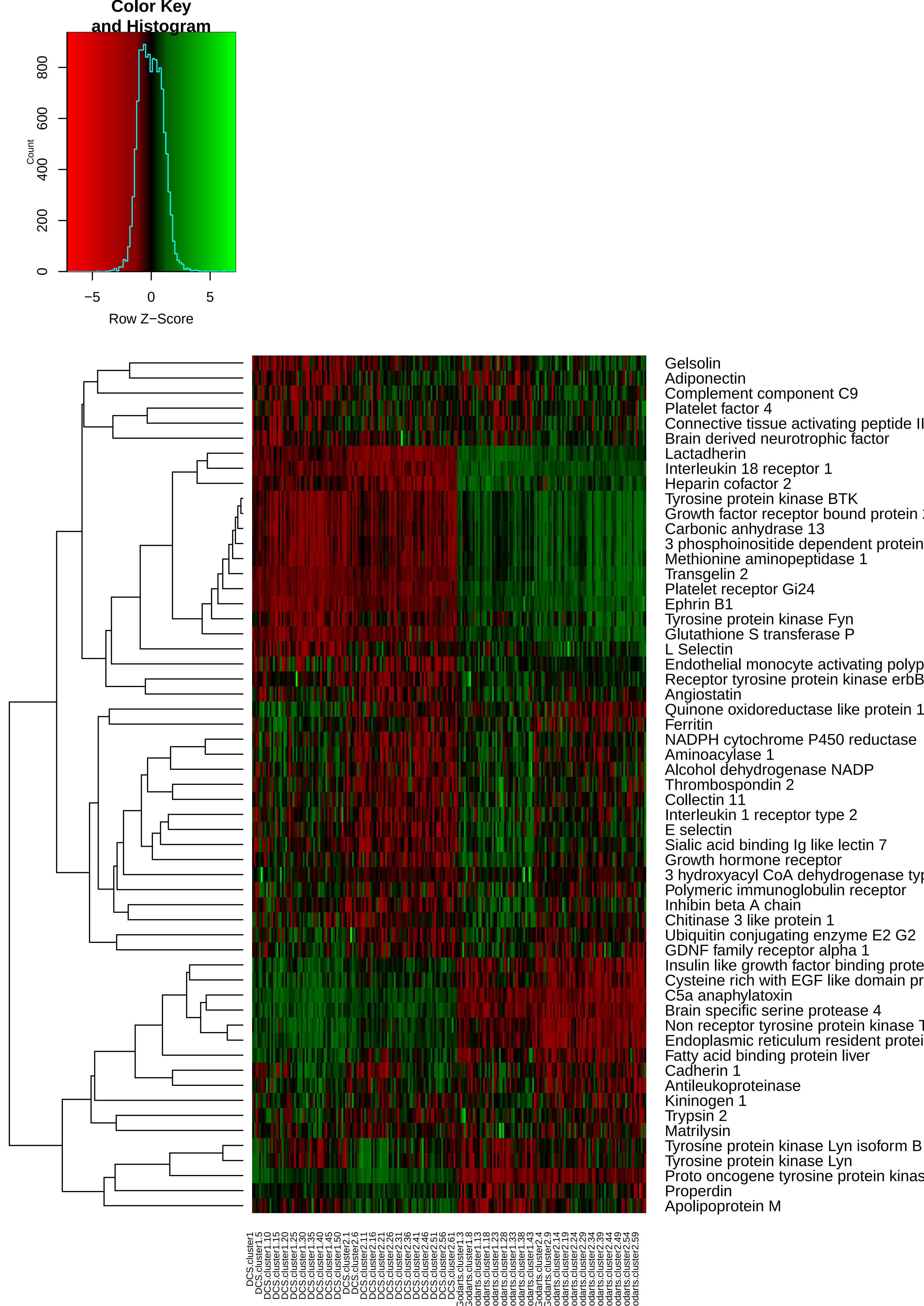

Color Key
and Histogram
Count
Row Z−Score
Gelsolin
Adiponectin
Complement component C9
Platelet factor 4
Connective tissue activating peptide II
Brain derived neurotrophic factor
Lactadherin
Interleukin 18 receptor 1
Heparin cofactor 2
Tyrosine protein kinase BTK
Growth factor receptor bound protein
Carbonic anhydrase 13
3 phosphoinositide dependent protein
Methionine aminopeptidase 1
Transgelin 2
Platelet receptor Gi24
Ephrin B1
Tyrosine protein kinase Fyn
Glutathione S transferase P
L Selectin
Endothelial monocyte activating polyp
Receptor tyrosine protein kinase erbB
Angiostatin
Quinone oxidoreductase like protein 1
Ferritin
NADPH cytochrome P450 reductase
Aminoacylase 1
Alcohol dehydrogenase NADP
Thrombospondin 2
Collectin 11
Interleukin 1 receptor type 2
E selectin
Sialic acid binding Ig like lectin 7
Growth hormone receptor
3 hydroxyacyl CoA dehydrogenase typ
Polymeric immunoglobulin receptor
Inhibin beta A chain
Chitinase 3 like protein 1
Ubiquitin conjugating enzyme E2 G2
GDNF family receptor alpha 1
Insulin like growth factor binding prote
Cysteine rich with EGF like domain pr
C5a anaphylatoxin
Brain specific serine protease 4
Non receptor tyrosine protein kinase T
Endoplasmic reticulum resident protei
Fatty acid binding protein liver
Cadherin 1
Antileukoproteinase
Kininogen 1
Trypsin 2
Matrilysin
Tyrosine protein kinase Lyn isoform B
Tyrosine protein kinase Lyn
Proto oncogene tyrosine protein kinas
Properdin
Apolipoprotein M

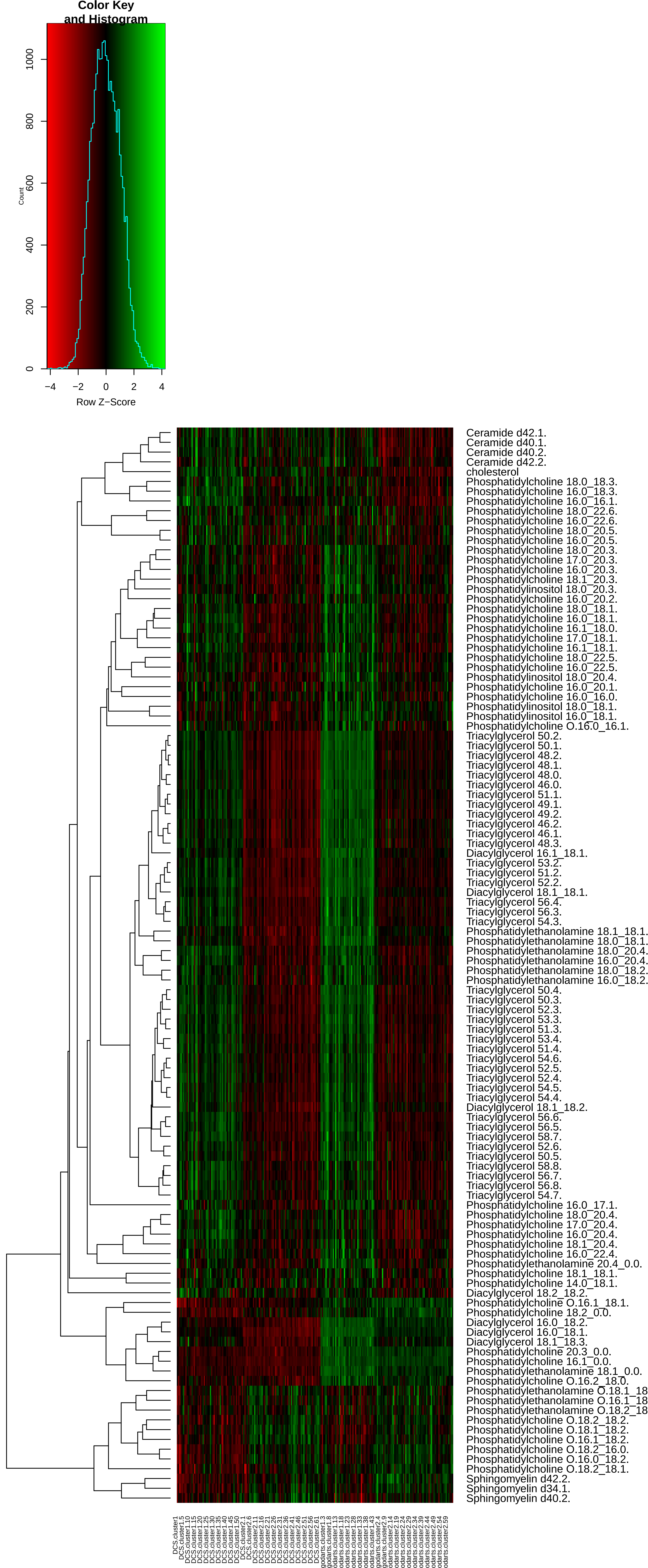
Color Key
and Histogram
Count
0
200
400
600
800
1000
−4
−2
0
2
4
Row Z−Score
Ceramide d42.1.
Ceramide d40.1.
Ceramide d40.2.
Ceramide d42.2.
cholesterol
Phosphatidylcholine 18.0_18.3.
Phosphatidylcholine 16.0_18.3.
Phosphatidylcholine 16.0_16.1.
Phosphatidylcholine 18.0_22.6.
Phosphatidylcholine 16.0_22.6.
Phosphatidylcholine 18.0_20.5.
Phosphatidylcholine 16.0_20.5.
Phosphatidylcholine 18.0_20.3.
Phosphatidylcholine 17.0_20.3.
Phosphatidylcholine 16.0_20.3.
Phosphatidylcholine 18.1_20.3.
Phosphatidylinositol 18.0_20.3.
Phosphatidylcholine 16.0_20.2.
Phosphatidylcholine 18.0_18.1.
Phosphatidylcholine 16.0_18.1.
Phosphatidylcholine 16.1_18.0.
Phosphatidylcholine 17.0_18.1.
Phosphatidylcholine 16.1_18.1.
Phosphatidylcholine 18.0_22.5.
Phosphatidylcholine 16.0_22.5.
Phosphatidylinositol 18.0_20.4.
Phosphatidylcholine 16.0_20.1.
Phosphatidylcholine 16.0_16.0.
Phosphatidylinositol 18.0_18.1.
Phosphatidylinositol 16.0_18.1.
Phosphatidylcholine O.16.0_16.1.
Triacylglycerol 50.2.
Triacylglycerol 50.1.
Triacylglycerol 48.2.
Triacylglycerol 48.1.
Triacylglycerol 48.0.
Triacylglycerol 46.0.
Triacylglycerol 51.1.
Triacylglycerol 49.1.
Triacylglycerol 49.2.
Triacylglycerol 46.2.
Triacylglycerol 46.1.
Triacylglycerol 48.3.
Diacylglycerol 16.1_18.1.
Triacylglycerol 53.2.
Triacylglycerol 51.2.
Triacylglycerol 52.2.
Diacylglycerol 18.1_18.1.
Triacylglycerol 56.4.
Triacylglycerol 56.3.
Triacylglycerol 54.3.
Phosphatidylethanolamine 18.1_18.1.
Phosphatidylethanolamine 18.0_18.1.
Phosphatidylethanolamine 18.0_20.4.
Phosphatidylethanolamine 16.0_20.4.
Phosphatidylethanolamine 18.0_18.2.
Phosphatidylethanolamine 16.0_18.2.
Triacylglycerol 50.4.
Triacylglycerol 50.3.
Triacylglycerol 52.3.
Triacylglycerol 53.3.
Triacylglycerol 51.3.
Triacylglycerol 53.4.
Triacylglycerol 51.4.
Triacylglycerol 54.6.
Triacylglycerol 52.5.
Triacylglycerol 52.4.
Triacylglycerol 54.5.
Triacylglycerol 54.4.
Diacylglycerol 18.1_18.2.
Triacylglycerol 56.6.
Triacylglycerol 56.5.
Triacylglycerol 58.7.
Triacylglycerol 52.6.
Triacylglycerol 50.5.
Triacylglycerol 58.8.
Triacylglycerol 56.7.
Triacylglycerol 56.8.
Triacylglycerol 54.7.
Phosphatidylcholine 16.0_17.1.
Phosphatidylcholine 18.0_20.4.
Phosphatidylcholine 17.0_20.4.
Phosphatidylcholine 16.0_20.4.
Phosphatidylcholine 18.1_20.4.
Phosphatidylcholine 16.0_22.4.
Phosphatidylethanolamine 20.4_0.0.
Phosphatidylcholine 18.1_18.1.
Phosphatidylcholine 14.0_18.1.
Diacylglycerol 18.2_18.2.
Phosphatidylcholine O.16.1_18.1.
Phosphatidylcholine 18.2_0.0.
Diacylglycerol 16.0_18.2.
Diacylglycerol 16.0_18.1.
Diacylglycerol 18.1_18.3.
Phosphatidylcholine 20.3_0.0.
Phosphatidylcholine 16.1_0.0.
Phosphatidylethanolamine 18.1_0.0.
Phosphatidylcholine O.16.2_18.0.
Phosphatidylethanolamine O.18.1_18
Phosphatidylethanolamine O.16.1_18
Phosphatidylethanolamine O.18.2_18
Phosphatidylcholine O.18.2_18.2.
Phosphatidylcholine O.18.1_18.2.
Phosphatidylcholine O.16.1_18.2.
Phosphatidylcholine O.18.2_16.0.
Phosphatidylcholine O.16.0_18.2.
Phosphatidylcholine O.18.2_18.1.
Sphingomyelin d42.2.
Sphingomyelin d34.1.
Sphingomyelin d40.2.

**Supplemental Figure 5. Features (lipids and proteins) that are strongly associated (P- value<=0.05) with SNF clusters in both DCS and GoDARTS cohorts, visualized as heatmaps.** Multi-omics clusters were generated from integrated lipidomics and peptidomics. Features association levels with each cluster were assessed by logistic regression and adjusted by gender, BMI and age. Concentrations of each feature were first converted into the log(concentration) and then normalized in order to investigate the relative expression level of each feature between different clusters (Row Z-score). Heatmap cells were coloured in red/green to represent the relative expression level (red=low, green=high). The federated database does not allow to pull individual sample's data in order to protect the patient's identity. Each vertical line in the heatmap represents 5 different patients' mean values for each feature.

Supplemental Table 1

| Clinical measures \ Pseudo R^2 | DCS | GoDARTS |
|---|---|---|
| Age | 0.0057 | 0.0053 |
| BMI | 0.0427 | 0.0097 |
| C-peptide | 0.0836 | 0.0303 |
| LDL Cholesterol | 0.0103 | 0.1167 |
| HDL Cholesterol | 0.0776 | 0.0717 |
| Triglycerides | 0.2589 | 0.4309 |
| Age+BMI+C-peptide+LDL Cholesterol+ HDL Cholesterol+Triglycerides | 0.2993 | 0.4689 |

Supplemental Table 1. Pseudo R square of six clinical measurements act as univariate or together as covariates for improvement of the prediction of multi-omics clustering assignments in a logistic regression model. Patients' multi-omics clustering assignments were treated as dependent variables and labelled as 1 or 0 (cluster #1:1, cluster #2:0). Pseudo R square was calculated based on the formula: Pseudo R square = 1-(deviance/null deviance).
